# Increased serum peripheral C-reactive protein is associated with reduced small-molecule brain perfusion in healthy volunteers and subjects with major depressive disorder

**DOI:** 10.1101/2020.06.29.20138438

**Authors:** Federico E. Turkheimer, Noha Althubaity, Julia Schubert, Maria A. Nettis, Oliver Cousins, Danai Dima, Valeria Mondelli, Edward T. Bullmore, Carmine Pariante, Mattia Veronese

**Affiliations:** Department of Neuroimaging, Institute of Psychiatry, Psychology and Neuroscience, King’s College London, London, UK; Department of Psychology, School of Arts and Social Sciences, City, University of London, London, UK; Department of Psychiatry, School of Clinical Medicine, University of Cambridge, UK; Cambridgeshire and Peterborough NHS Foundation Trust, Cambridge, UK; Immuno-Psychiatry, Immuno-Inflammation Therapeutic Area Unit, GlaxoSmithKline R&D, Stevenage, UK

**Keywords:** inflammation, depression, BBB permeability, CSF, choroid plexus, PET, tracer percolation

## Abstract

The relationship between peripheral and central immunity and how these ultimately may cause depressed behaviour has been the focus of a number of imaging studies conducted with Positron Emission Tomography (PET). These studies aimed at testing the immune-mediated model of depression that proposes a direct effect of peripheral cytokines and immune cells on the brain to elicit a neuroinflammatory response via a leaky blood-brain barrier and ultimately depressive behaviour. However, studies conducted so far using PET radioligands targeting the neuroinflammatory marker 18 kDa translocator protein (TSPO) in patient cohorts with depression have demonstrated mild inflammatory brain status but no correlation between central and peripheral immunity.

To gain a better insight into the relationship between heightened peripheral immunity and neuroinflammation, we estimated blood-to-brain and blood-to-CSF perfusion rates for two TSPO radiotracers collected in two separate studies, one large cross-sectional study of neuroinflammation in normal and depressed cohorts and a second study where peripheral inflammation in healthy controls was induced via subcutaneous injection of interferon (IFN)-α. In both studies we observed a consistent negative association between peripheral inflammation, measured with c-reactive protein P (CRP), and radiotracer perfusion into and from the brain parenchyma and CSF. Importantly, there was no association of this effect with the marker of BBB leakage S100β, that was unchanged, but there was an association between the reduction of tracer perfusion in volunteers injected with interferon (IFN)-α and VEGF, a potent vascular permeability factor.

These results support a different model of peripheral-to-central immunity interaction whereas peripheral inflammation causes a “stiffening” of the healthy BBB with consequent reduction of small molecule trafficking to and from the blood into the brain and CSF. This effect, on the long term, is likely to disrupt brain homeostasis and induce depressive behavioural symptoms. Moreover, given the molecular similarity between the TSPO ligands and antidepressant, this phenomenon may underlie treatment resistance in depressive cohorts with heightened peripheral status.

## INTRODUCTION

The relationship between peripheral and central inflammation and depressed behaviour has been investigated by a number of in vivo studies using Positron Emission Tomography (PET) and radioligands targeting the 18 kDa translocator protein (TSPO) in patients with depression. The TSPO is a mitochondrial protein that is expressed in a number of cells (endothelial, astrocyte) of the central nervous system (CNS) but is particularly enriched in activated microglia, the brain-resident macrophages (Betlazar et al., 2018; Tournier et al., 2019). However, TSPO PET studies in depressed cohorts have returned mixed results demonstrating either negative (Zhang et al., 2018), null (Hannestad et al., 2013) or mild TSPO elevations (Holmes et al., 2018; Richards et al., 2018; Setiawan et al., 2015; Su et al., 2016; Zhang et al., 2018), with elevation being more evident in unmedicated subjects (Richards et al., 2018) and in those with suicidal thoughts (Holmes et al., 2018). More importantly, so far no study has demonstrated a consistent and replicable association between TSPO brain concentration and peripheral inflammatory mediators (Enache et al., 2019; Mondelli et al., 2017). A positive association was found in a sample of 48 patients with major depressive disorder between plasma adiponectin (an anti-inflammatory protein) and TSPO PET imaging in anterior cingulate cortex, but the analysis was only exploratory and did not survive correction for multiple comparisons (Richards et al., 2018). More recently, a negative association between CRP and brain TSPO expression was found by Attwells (Attwells et al., 2019), suggesting that patients with depression and high peripheral inflammation would have less microglia activation than those with normal level blood CRP. This disagrees with the immune-mediated model of depression for which depressive symptoms can be induced by peripheral cytokines and immune cells acting on the brain to elicit a neuroinflammatory response via a leaky blood-brain barrier (BBB) (Dantzer, 2009; Miller and Raison, 2016; Schedlowski et al., 2014). The evidence above prompted us to re-evaluate the data collected in TSPO imaging studies in normal and depressed cohorts by focusing on the relationship with one particular peripheral inflammatory marker, C-reactive protein (CRP), in order to propose an alternative model for the mechanism of action between peripheral and CNS immunity.

Meta-analysis of cross-sectional and longitudinal studies on the relationship between peripheral immunity and depression have highlighted the relevance of pro-inflammatory cytokines such as tumour necrosis factor-α (TNF-α), interleukin-1β (IL-1β) and interleukin-6 (IL-6) but also of the acute phase protein, CRP (Valkanova et al., 2013). CRP is an acute phase reactant, a protein primarily made by the liver that is released into the blood within a few hours after tissue injury, the start of an infection or other inflammatory insults (Sproston and Ashworth, 2018). The concentration of CRP in blood has been increasingly used to quantify levels of systemic inflammation and associated levels of risks for a number of conditions, with concentrations greater than 3 mg/L indicating a high risk of cardiovascular disease (Smith, 2004).

In-vitro and preclinical models have shown that high concentrations of CRP (10 to 20 mg/L) are associated with BBB disruption, increasing its permeability (Hsuchou et al., 2012). However, the clinical relevance of this literature to the in-vivo human context remains to be shown, as in most instances CRP levels in depressed cohorts are below the 10 mg/L mark, which is on the low to moderate spectrum of the peripheral inflammatory range (Elwood et al., 2017); in particular, we note that higher CRP levels increase BBB porosity to plasma proteins (measured as cerebrospinal fluid/serum albumin ratio) but only in those instances when CSF also exhibited abnormalities (Elwood et al., 2017). At the same time, other in-vitro work has reported that milder CRP plasma concentrations may stiffen endothelial cells and reduce endothelial permeability (Kusche-Vihrog et al., 2011; Tomiyama et al., 2005). Importantly, a very recent epidemiological analysis on a sample of ∼120,000 adults has shown that, together with metabolic syndrome, the increased level of CRP explained 37% of the association between worse arterial stiffness and depression (Dregan et al., 2020).

Hence, we hypothesized that, in normal volunteers and patients with depression but no other BBB impairment, the levels of CRP in the range observed in these cohorts (range 3-10 mg/l, equivalent to ‘low-grade inflammation’) are associated with reduced blood to brain permeability. To test this hypothesis, we evaluated previously acquired data on the perfusion of TSPO radiotracers across the blood-brain interface (e.g., the BBB) and the blood-cerebrospinal fluid (CSF) interface (e.g., the choroid plexus).

Specifically, we quantified radiotracer perfusion from blood into parenchyma (via the BBB) and from blood and parenchyma into the CSF (via epithelial barriers and the choroid plexus) from TSPO PET data collected in two separate studies, one large cross-sectional study of neuroinflammation in normal controls and depressed patients (Schubert et al., 2020) and a second study where peripheral inflammation in normal volunteers was induced via subcutaneous injection of interferon (IFN)-α and CNS inflammation was measured with PET (Nettis et al., 2020).

## METHODS

### Datasets

Two independent TSPO PET datasets collected from two separate neuroimaging studies were considered in this work.

*Dataset* 1 includes 51 subjects with major depressive disorder (MDD) (36/15 women/men; mean age: 36.2 ± 7.4 years) and 25 matched healthy controls (HC) (14/11 women/men; mean age: 37.3 ± 7.8 years) as part of the Biomarkers in Depression Study (BIODEP, NIMA consortium, https://www.neuroimmunology.org.uk/) to investigate the contribution of neuroinflammation in depression. Patients aged 25 to 50 (inclusive) that were taking anti-depressant treatment and had a total Hamilton Depression (HAMD) score>13 or were untreated and had a total HAMD>17 were included. The final sample consisted of 9 untreated patients and 42 taking medications. MDD patients were further classified based on their blood CRP level, CRP >3mg/L corresponding to the high CRP group (20 patients, 36.9 ± 7.6 years) and CRP<=3mg/L corresponding to *low CRP group* (31 patients, 35.7 ± 7.2 years). CRP concentrations for controls were no different to the ones for low CRP group (t =0.017, p=0.99). MDD cases and HC with a lifetime history of other neurological disorders, active drug and/or alcohol abuse, participation in clinical drug trials within the previous year, concurrent medication or medical disorder that could compromise the interpretation of results, and those who were pregnant or breastfeeding were excluded. HC subjects had no personal history of clinical depression requiring treatment and were age- and sex-matched with the patient group. On the contrary, BMI was significantly higher for the depression group (BMI_MDD_: 27.2±4.0) kg/m; BMI_HC_ 24.2±4.8 kg/m^2^; p-value=0.001).

*Dataset* 2 includes data from 7 healthy males to image microglia activation in the CNS before and ∼24 hours after one subcutaneous injection of the immune challenge IFN-α 2a (Roferon-A 3 million IU/0.5 ml solution for injection). The study aimed to investigate putative changes in brain microglia activity and their relationship with changes in peripheral inflammation and in mood. Eligible participants were non-smokers, drank no more than 5 alcohol drinks per week, had no history of significant medical illness and did not meet the criteria for any current or past psychiatric or substance-dependence diagnosis. Subjects were excluded if they had an infection in the last month or had regularly used anti-inflammatory drugs. All subjects were TSPO high-affinity binders (HABs) as determined by single-nucleotide polymorphism rs6971 genotyping (Owen et al., 2012).

All imaging protocols were approved by the local ethics committee and participants gave written informed consent prior to data collection.

### Image acquisition and analysis

*Dataset 1* - All subjects underwent 60-minute dynamic PET scan on a GE SIGNA PET/MR scanner (GE healthcare, Waukesha, USA) after an intravenous bolus injection of [^11^C]PK11195 (target dose ∼350 MBq, injected dose 361±53MBq). Data were reconstructed using multi-subject atlas method and improvements for the MRI brain coil component (Burgos et al., 2014). Data were corrected for scatter, randoms and deadtime using the scanner GE software. No arterial blood data was collected during the PET imaging consistent with best practice of [^11^C]PK11195 PET imaging (Turkheimer et al., 2007). Full experimental details are reported in the original reference (Schubert et al., 2020).

*Dataset 2* – All the subjects underwent two 90-minute dynamic PET scans on a Siemens Biograph™ True Point™ PET/CT scanner (Siemens Medical Systems, Germany) after receiving a bolus injection of [^11^C]PBR28 (target dose ∼350 Mbq, injected dose 341±15). Images were reconstructed using filtered back projection and corrected tissue attenuation, scatter and random coincidences, and deadtime using scanner software. In parallel to the PET acquisition, arterial blood sampling was performed to generate metabolite-free arterial input function consistently with previous studies using the same tracer (Bloomfield et al., 2016). Full experimental details are reported in the original reference (Nettis et al., 2020).

In both datasets, all subjects also had structural T1-weighted brain MRI, either collected during the PET acquisition as in *dataset 1* or acquired just before the PET session as in *dataset 2*. Tehse MRI images were used for PET data processing including grey matter (GM) and white matter (WM) tissue segmentation, brain masking, and atlas-based region extraction. These pre-processing steps as well as the PET inter-frame motion correction were performed using MIAKAT™ (www.miakat.org) a MATLAB-based software combining tools from SPM12 and FSL for PET/MRI imaging analysis. For the [^11^C]PK11195 PET studies TSPO density was assessed using BPnd parameter calculated by using a supervised clustering reference region approach (Turkheimer et al., 2007). For the [^11^C]PBR28 PET study, instead, the tracer tissue kinetics was described with full compartmental modelling (Rizzo et al., 2019; Rizzo et al., 2014). The tracer blood to tissue exchange rate (*K*_*1*_) and tracer volume of distribution (V_T_) were used as main parameters of interest to measure the tracer perfusion and TSPO availability, respectively.

### Quantification of blood to CSF tracer exchange

TSPO tracer exchange between blood and CSF through CP was calculated from the PET imaging using the validated analysis protocol defined in (de Leon et al., 2017; Schubert et al., 2019). The method has shown to be sensitive to CSF dynamics, returning evidence for altered CSF-mediated clearance in dementia and multiple sclerosis (de Leon et al., 2017; Schubert et al., 2019). Briefly, manual lateral ventricle ROIs were generated for all subjects using the subject T1-weighted structural MRI data and the ITK-SNAP (Yushkevich et al., 2006) (itksnap.org) snake tool, following previously described guidelines for lateral ventricle extraction (Acabchuk et al., 2015). The lateral ventricle ROIs were then eroded by two voxels (5.2 mm) using the erode function given by FSL’s ‘fslmaths’ utility package in order to reduce partial volume effects of the surrounding tissues.

For both datasets, standardized uptake value ratios (SUVRs) at 60 minutes and the area-under-curve from 30 to 60 minutes (AUC30-60) were calculated from the PET images using the eroded lateral ventricle activity as target region and supervised reference region (dataset 1) or whole brain gray matter (dataset 2) as normative region.

In dataset 2, for which metabolite-free arterial input function was also available, we implemented the compartmental modelling approach to quantitatively describe the rates of exchanges of the tracer from blood to lateral ventricle (*K1*), from tissue into lateral ventricle and clearance (*kClearance*) as well as the tracer specific biding in the lateral ventricle (*kon* and *koff*) (Figure 1). The volume of distribution of the [11C]PBR28 into lateral ventricles was calculate as V_T_ = *K*_*1*_/kClearance*(1+kon/koff). Interestingly, it was not possible to model a specific tracer exchange from tissue to CSF as in the original paper describing [11C]PIB kinetics into lateral ventricles (Schubert et al., 2019); this may be possibly due to a poorer extraction of the radiotracer (Sander et al., 2019). Consistent with a previous publication the model was implemented using SAAM II software (Barrett et al., 1998).

**Figure 1.**
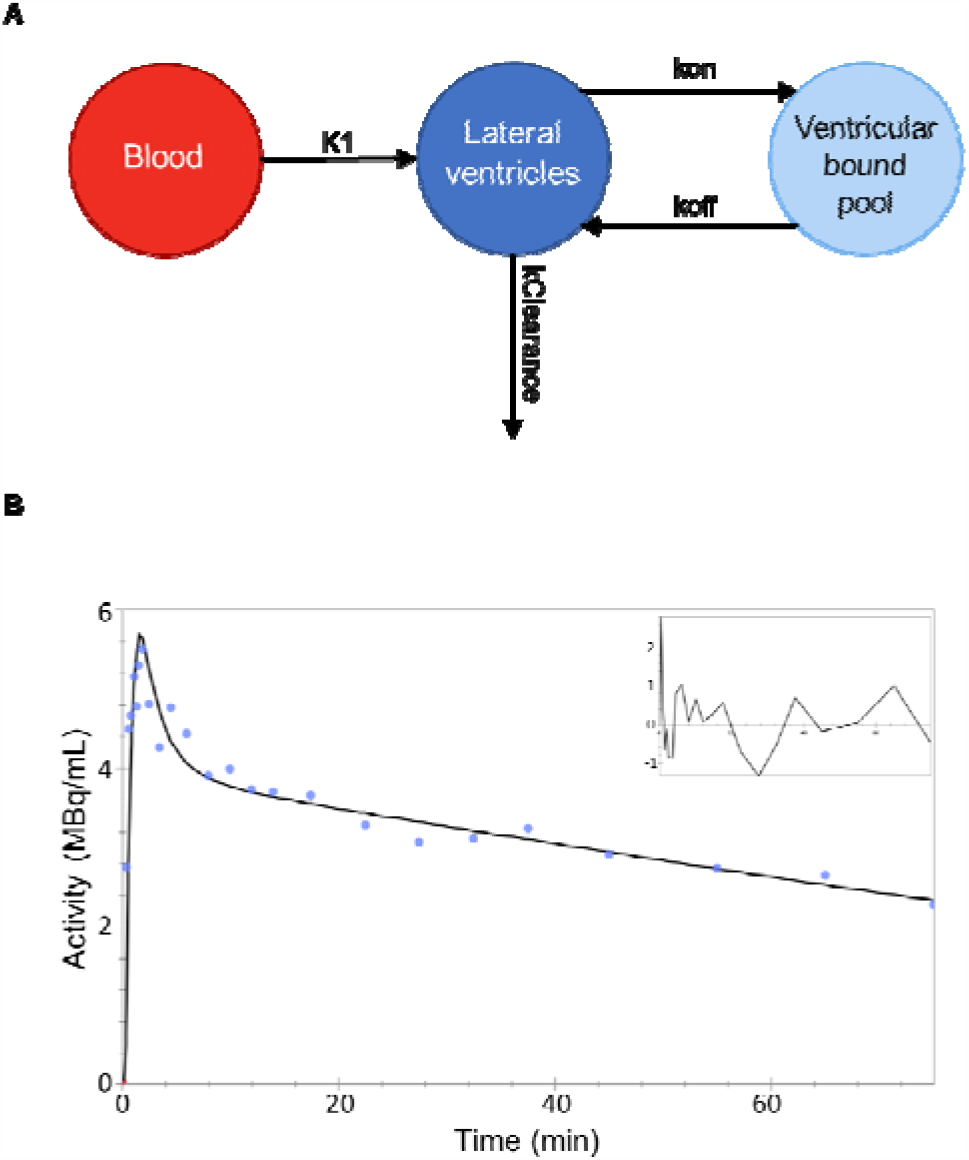
Compartmental modelling of [11C]PBR28 tracer kinetic in lateral ventricles. *A) Compartmental model and kinetic parameters. B) Representative data fit (healthy control, baseline)*.

### High sensitivity C-reactive protein measurement

For both datasets venous blood samples were collected from each participant for the CRP analysis In study 1, a venous blood sample was collected after an overnight fast between 08:00 and 10:00 on the day of clinical assessment. Participants had refrained from exercise for 72 h and had been lying supine for 0.5 h prior to venipuncture (Chamberlain et al., 2019). For study 2, blood samples were collected at the time of PET scans, i.e. at baseline and ∼24 hours after IFN-α injection. High sensitivity CRP (hsCRP) was used to assess peripheral inflammation. For both projects, blood samples were collected in clot activator containing tubes for measurement. The samples were allowed to coagulate for 30–60 minutes, then centrifuged at 1600 Relative Centrifugal Force (RCF) for 15 min. For the first project (BIODEP), serum samples were separated and transported to a central laboratory (Q2Solutions) where analyzed on the day of receipt. Samples were exposed to anti-CRP-antibodies on latex particles, and the increase in light absorption due to complex formation was used to quantify hsCRP levels, using Turbidimetry on Beckman Coulter AU analyzers. Inter and intra-assay co-efficient of variations were<10%. For the second project (FLAME) after blood collection, the serum was separated, aliquoted and stored at −80 °C before use. It was later assayed on the Siemens Advia 2400 Chemistry analyser (Siemens Healthcare Diagnostics, Frimley, UK) (Mason et al., 2013).

Forth both datasets, VEGF-A was measured using Meso Scale Discovery (MSD) V-PLEX sandwich immunoassays (Dabitao et al., 2011) (King et al., 2019), and plates read on an MSD QuickPlex SQ 120. The results were analyzed using MSD DISCOVERY WORKBENCH analysis software. In dataset 2 (FLAME), levels of serum S100B protein were also measured in serum using a S100B kit distributed by Diasorin, Charles House, Toutley Road, Wokingham, Berkshire, run on the Liaison XL chemiluminescence analyser (Townend et al., 2002; Wunderlich et al., 2004).

### Statistics

SPSS (version 24.0, Chicago, IL) was used to perform all statistical analyses. Normality of the data was tested using Shapiro-Wilk’s W test. For *dataset 1*, blood to CSF exchange measures (i.e. SUVR and AUC30-60) were investigated using analysis of variance (ANOVA) to test for group differences between MDD at different CRP levels and HC, while covarying for possible confounding factors (e.g. age, BMI, tracer injected dose, or lateral ventricle volume). Group differences were further investigated using independent samples t-tests, while association between CSF-mediated clearance measures and clinical scores (i.e. HAMDI, Childhood trauma, and Perceived Stress Score) were investigated with bivariate correlations. Correlation of these metrics with the blood brain barrier permeability marker (VEGF) was also explored. For *Dataset 2*, we explored the relationship between CRP changes and blood to CSF exchange measures (i.e. SUVR and AUC30-60) before and after IFN-α challenge. We also compared *K*_*1*_ and V_T_, both into lateral ventricles and GM parenchyma between conditions. The latter was chosen as representation of the parenchymal tracer uptake independent from lateral ventricles. Correlation of these metrics with CRP levels, peripheral tracer plasma protein binding and the blood brain barrier integrity markers (VEGF and S100β) were also explored. We implemented the Durbin-Watson test to exclude outliers.

## RESULTS

### Marked inverse association between blood-to-CSF exchange measures and CRP in patients with depression

Results from the analysis of dataset 1 indicated the presence of a reduction in blood to CSF small-molecule perfusion as shown by an inverse association between blood-to-CSF exchange parameters and CRP, with higher CRP levels corresponding to both lower SUVR and lower AUC30-60 (Figure 2) in both MDD patients and HC. Considering the whole population (HC and MDD cases) the association remained significant, once corrected for lateral ventricle volumes using partial correlations, for both SUVR (r=-0.233, p=0.045) and AUC30-60 (r=-0.277, p=0.016). There was no effect of age, tracer injected dose and subject BMI on SUVR and AUC30-60 (p>0.4).

These associations between CRP and CSF radiotracer activity became stronger when considering MDD patients only, both in SUVR (partial correlation r: −0.300, p=0.034) and AUC30-60 (partial correlation r: −0.338, p=0.016). The relationship between these variables became more evident by restricting the analysis to the MDD-high CRP group only. Controlling for lateral ventricle volume SUVR-CRP partial correlation was −0.447 (p=0.048) while AUC30-60-CRP partial correlation was −0.579 (p=0.007). As for the whole sample, there was no effect of age, tracer injected dose and subject BMI on SUVR and AUC30-60 correlation with CRP.

No correlations between CSF-mediated clearance measures and HAMD score, Childhood trauma or Perceived Stress Score were found. The only exception was a significant inverse association between AUC30-60 and Childhood trauma score (Spearman’s rho = −0.32, p: 0.02) although it did not survive to multiple comparison correction.

No correlation between VEGF and CSF-mediated clearance measures or CRP was found (p>0.5) although MDD cases reported a significantly lower VEGF value compared to HC (t_44_: 2.90, p<0.01).

**Figure 2.**
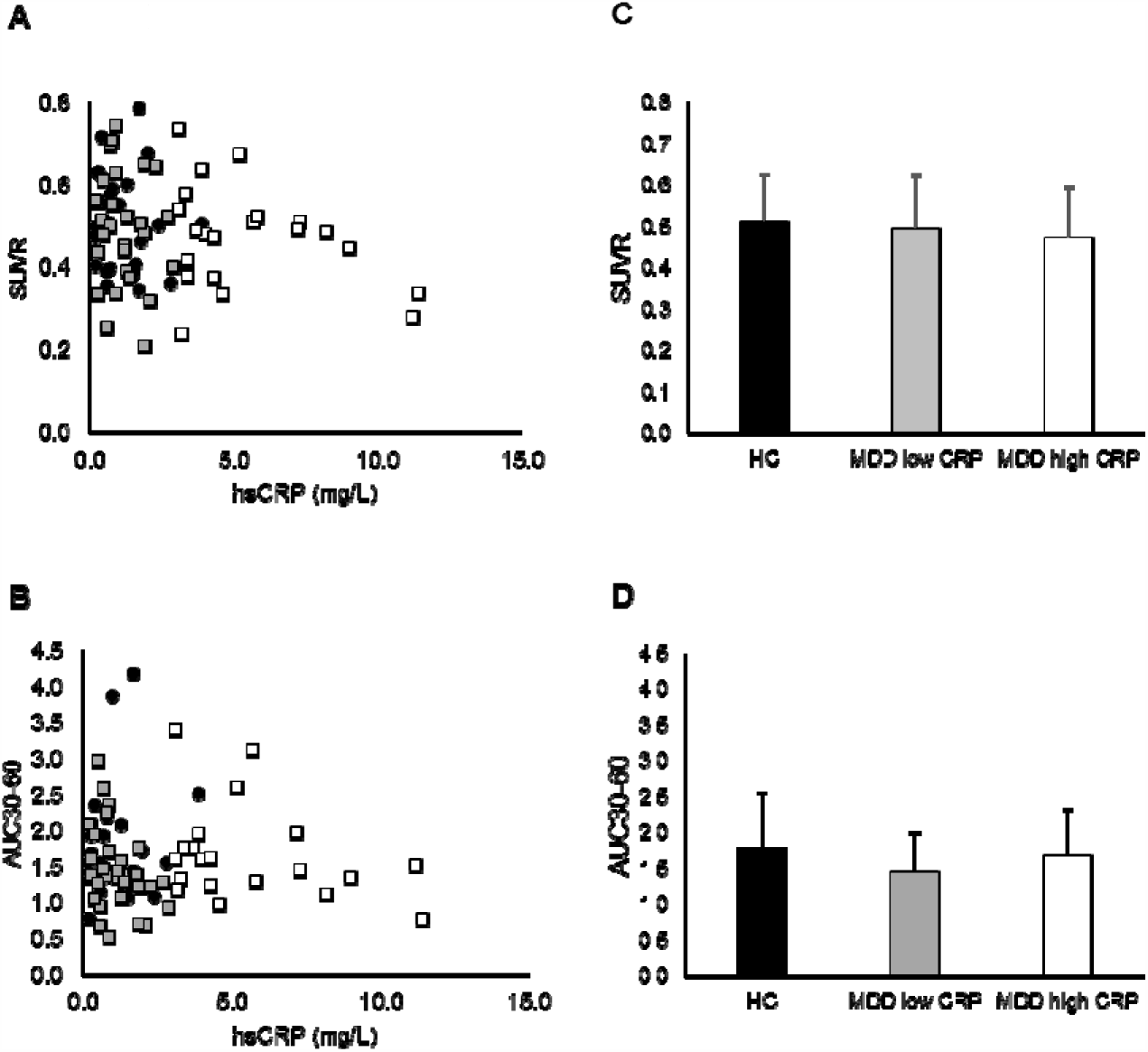
CSF-mediated clearance parameters and peripheral CRP in MDD. *The results refer to [11C]PK11195 PET study (dataset 1). Patients are identified by low CRP group (<3mg/L, grey bar) and high CRP group (>3mg/L, white bar). Healthy matched controls are reported in black. Panel A and B show the distribution of SUVR and AUC30-60 as function of CRP, respectively. Panel C and D show the SUVR and AUC30-60 group averages and standard deviations*.

### Peripheral CRP increase is associated with reduction of blood to brain tracer percolation after IFN-α injection

In dataset 2, we found that the CRP increase following the peripheral IFN-α challenge was inversely associated with lateral ventricular SUVR measure (R^2^: 0.72, p=0.015) (Figure 3A) indicating a reduction on blood-to-CSF small molecule transfer. This association was not replicated by AUC30-60 (Figure 3B).

The most interesting results were obtained for the compartmental modelling analysis (Figure 3C and D). When we considered the tracer kinetics into lateral ventricle CSF we found that V_T_ was significantly reduced after IFN-α injection (−18±12%, p=0.01) while the difference in blood to CSF tracer transportation *K*_*1*_ almost reached significance (−23±20%, p=0.05). No significant difference was found for the other kinetic parameters (kClearance: 1±49%, kon: 19±56%; koff: 2±10%).

**Figure 3.**
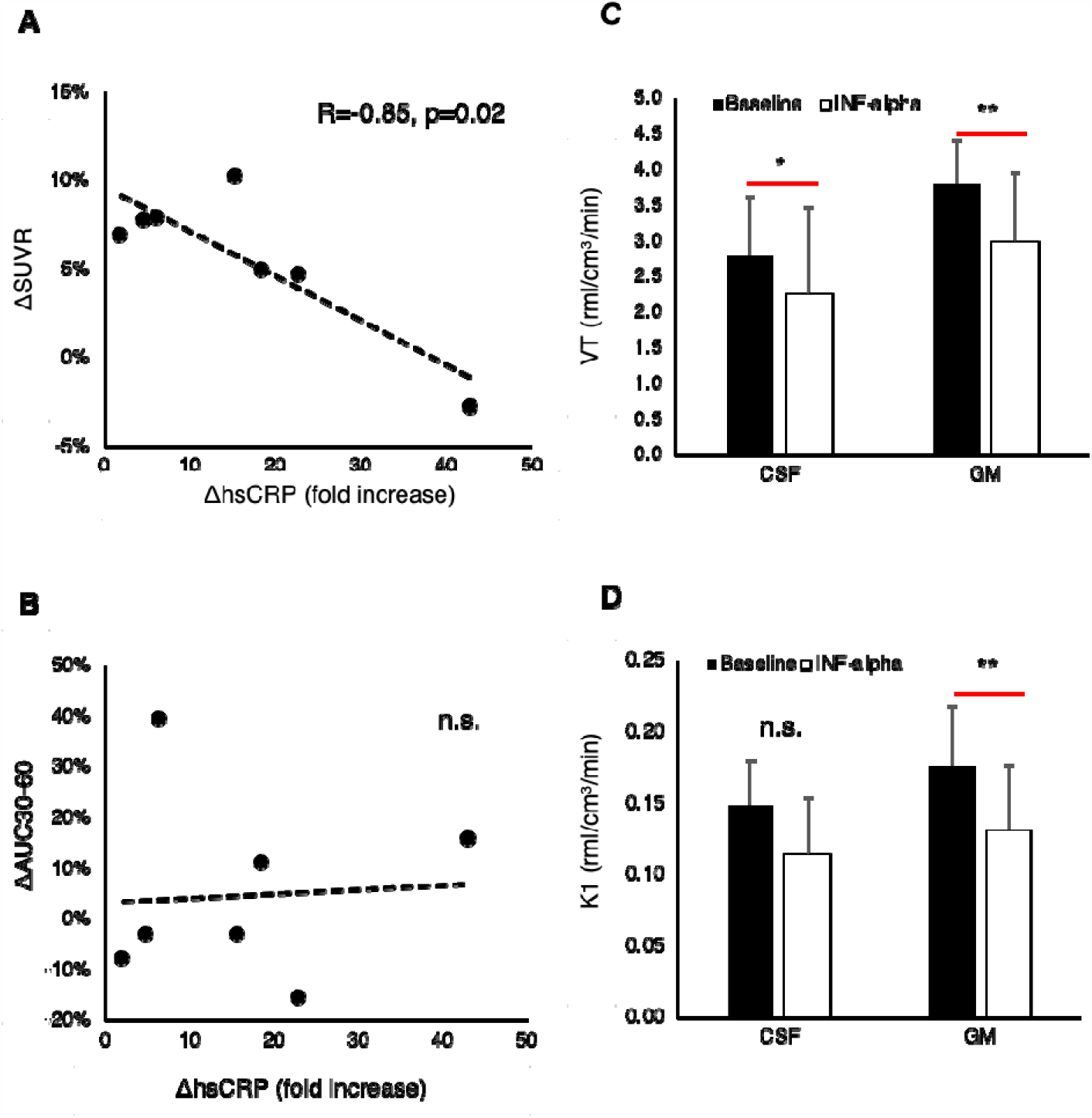
CSF-mediated clearance after IFN-α immune challenge. *The results refer to [11C]PBR28 PET study (dataset 2). A) Association between CRP and SUVR variations at baseline and after 24 hours IFN-α injection. B) Association between CRP and AUC30-60 variations at baseline and after 24 hours IFN-α injection. C) Comparison of the volume of distributions (VT) for GM and lateral ventricle CSF at baseline and after 24 hours IFN-α injection. D) Comparison of blood to tissue rates (K*_*1*_*) for GM and lateral ventricle CSF at baseline and after 24 hours IFN-α injection*.

A stronger effect of interferon-α was found also for the kinetic parameters describing tracer kinetics with brain GM. Particularly we found that both blood to GM tracer transportation *K*_*1*_ (−27±15%, p<0.01) and GM V_T_ (−22±10%, p<0.01) were significantly reduced after the challenge. Given the effect that blood proteins have on the TSPO tracer availability, we explored whether these reductions were associated with CRP changes or with tracer plasma protein binding (Figure 4). We found that both variables were associated with reduction of tracer percolation into GM (Figure 4A and B) and into lateral ventricle CSF (Figure 4D and E).

Given the collinearity between plasma protein binding and CRP (R^2^=0.32, p=0.03) we studied their interaction with *K_1_* as the dependent variable. Partial regression between K_1_ and CRP controlling for plasma protein binding (fp) showed significant association for GM tissue (Pearson’s R=-0.65,p<0.01) but not for CSF (Spearman rho=-0.37, p=0.11). However, when we directly corrected the transfer rate K_1_ by multiplication with fp, we found significant associations between the tracer perfusion and CRP for both CSF and GM (Figure 4C and F).

**Figure 4.**
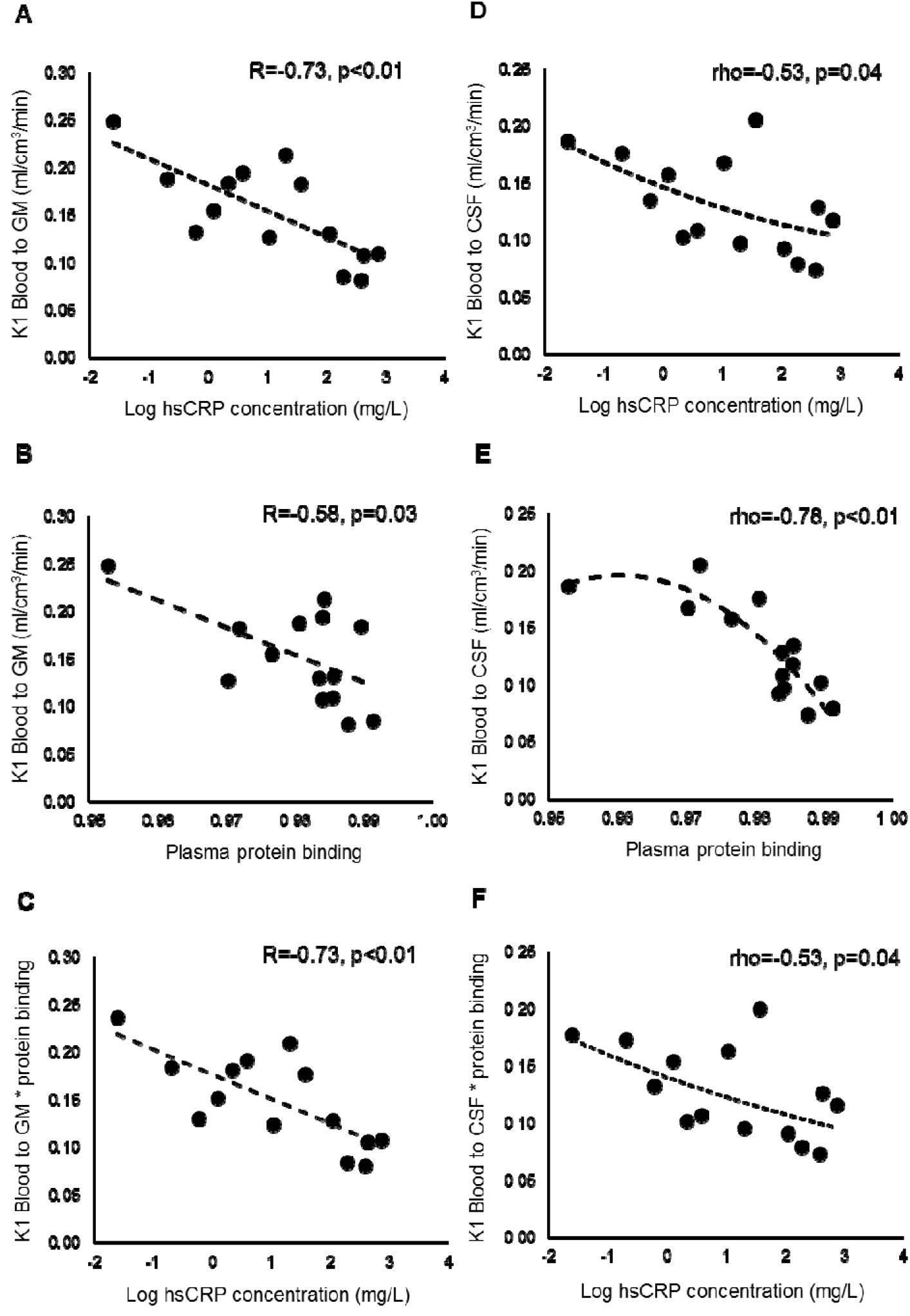
Association between blood to tissue tracer percolations, plasma protein binding and CRP. *A-C) Blood to tissue transport into brain GM. D-F) Blood to tissue transport into lateral ventricle CSF*.

In term of BBB permeability markers, there were positive associations between VEGF and *K*_*1*_ parameters (R=0.63, p=0.02 for CSF and R=0.53, p=0.05 for GM) but no association was found between S100β and any kinetic parameter (both *K_1_* and tracer clearance from lateral ventricle) (Figure 5). Interestingly S100β and VEGF were positively associated (R=0.55, p=0.04) but only VEGF was significantly reduced after interferon-α (from 134.5±52.5 at 1 h before IFN-α to 93.5±34.9 at 24 hours after; paired t-test, t=5.15, p=0.002).

**Figure 5.**
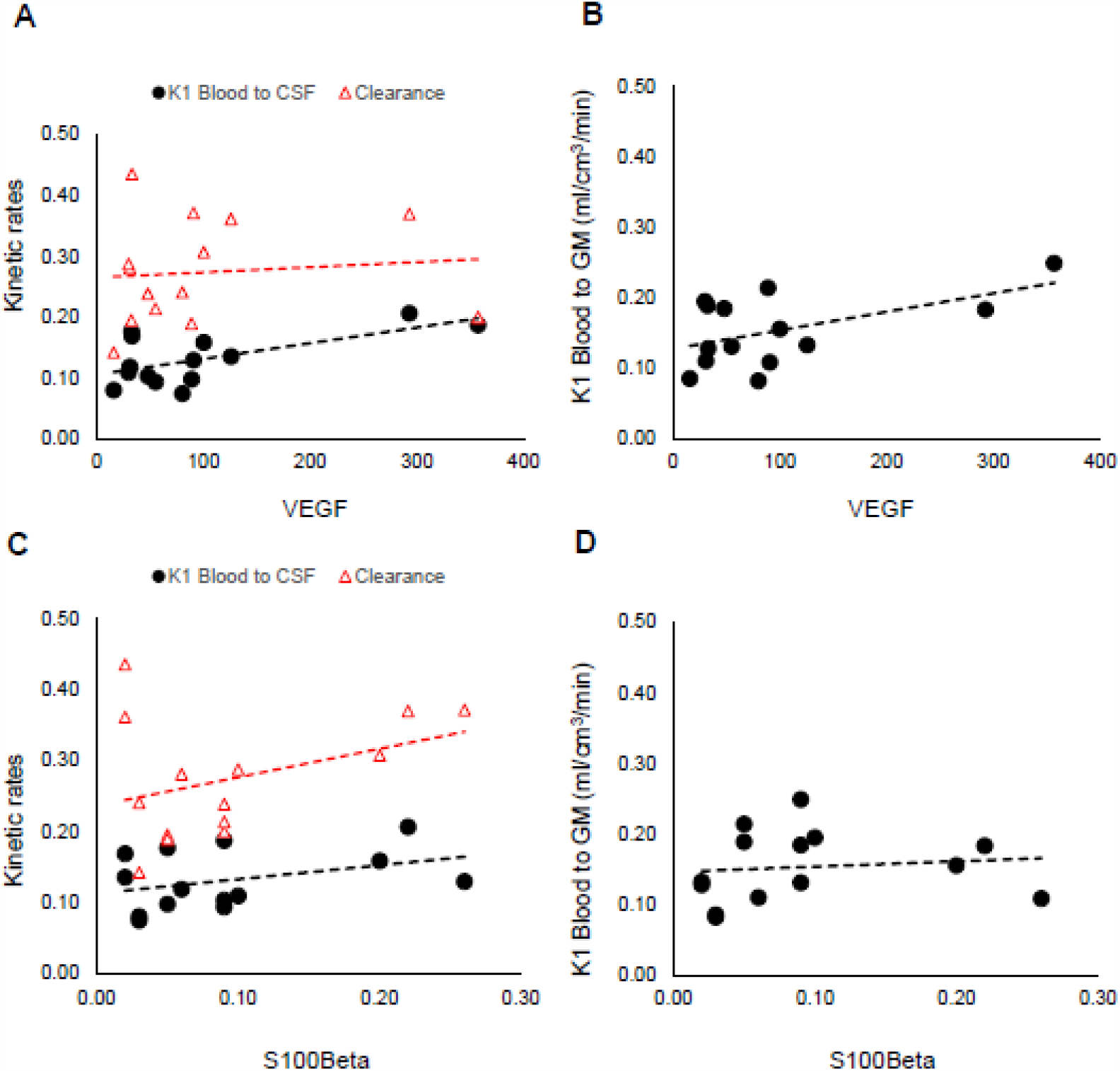
Association between blood to tissue tracer percolations and BBB integrity (VEGF and S100β). *A) Tracer kinetics in lateral ventricles (black circles = K_1_, red triangle = kClearance). B) Blood to GM transport (black circles = K_1_)*

## DISCUSSION

This work has demonstrated that high levels of the peripheral inflammatory marker CRP are associated with reduced influx of PET radiotracers into brain spaces. The implications of this study are that a reduction of brain/blood exchanges of small molecules is present in the conditions of activated peripheral immune activity, with possible downstream effects on brain homeostasis that are potentially relevant to the development of depression. In addition, on a technical level, our findings are relevant in the context of the difficulties encountered in the quantification of the binding of TSPO PET tracers in the study of patient cohorts with inflamed peripheral status, as in case of heightened peripheral inflammatory status, the volume of distribution, e.g. the estimated ratio between the target and the plasma tracer concentrations, will be lowered by the decreased perfusion and this may be mistakenly interpreted as a reduction in the target TSPO density. This further complicates the interpretation of TSPO neuroimaging studies, which is already methodological challenging (Turkheimer et al., 2015).

Depression’s main environmental risk factor is chronic stress and one prevalent model for this interaction is that peripheral myeloid cells or pro-inflammatory cytokines can diffuse into the brain of stressed individuals due to stress-induced increase of BBB permeability; a large body of literature exists on this matter (Esposito et al., 2001; Friedman et al., 1996; Hodes et al., 2015; Menard et al., 2017; Santha et al., 2015; Weber et al., 2017; Wohleb et al., 2013) although this link is still controversial (Roszkowski and Bohacek, 2016). Clinical evidence is very sporadic with only a few reports on the subject; association between BBB leakage and depression has been demonstrated with higher CSF albumin concentration in small patients subgroups (Bechter et al., 2010) and in elderly women (Gudmundsson et al., 2007). Another marker of BBB integrity is the astrocytic marker S100β protein that has been reported as increased in MDD (Arora et al., 2019; Tsai and Huang, 2016) and reduced after medication (Schroeter et al., 2002), although elevated levels S100β have been shown to be correlated positively with therapeutic response in depressed patients (Ambree et al., 2015; Arolt et al., 2003), with few exceptions (Tsai and Huang, 2016). Contrary to this, in MDD cohorts, high levels of CRP in plasma and CSF are not associated with increased BBB permeability as measured by albumin plasma-to-CSF ratio (Felger et al., 2018). Indeed, even in this paper, patients in the BIODEP cohort did not demonstrate biomarkers of BBB disruption; instead they did show a reduction of VEGF (a potent vascular permeability factor (Jiang et al., 2014)) that may indicate reduction in BBB permeability. In the FLAME study, the HC after the IFN-α challenge did not exhibit evidence of BBB leakage as measured by serum levels of S100β, but VEGF resulted significantly reduced. Similarly results have been found to modulate a brain regional propagation of peripheral inflammation by selective activation of BBB endothelial cells {Süß, 2020 #113}.

The data in this work instead suggest a possible different model that posits a reduction of the permeability of the healthy BBB in reaction to increased peripheral inflammation as indexed by CRP; this at least seems to be the case for small molecules, given the molecular weight of the tracers used (352.9 g/mol for PK11195 and 347.4 g/mol for PBR28). Hence, we may then speculate that the mild inflammatory state observed by PET imaging in the brain of clinically depressed patients may be the result of disturbed homeostasis due to the much-reduced transfer of solutes across the BBB. As confirmed by the IFN-alpha model in healthy humans, the observed reduction of *K*_*1*_ after the IFN-α challenge (up to a third) would impede the normal exchange of most lipid-soluble molecules as well as small polar metabolites that are ported passively through the barrier (Johansen et al., 2018). This reduction of BBB molecular transfer may be protective on the short-term but may lead to disturbed homeostasis in the case of prolonged/chronic inflammatory states. If this were proven to be the case, then the mechanism of interaction between peripheral cytokines and CNS immune activity would be indirect, e.g., mediated by the BBB closure and ensuing disturbance to CSF dynamics.

In MDD there is often increased peripheral circulating glucocorticoids (Lombardo et al., 2019), and conditions with high endogenous glucocorticoids, such as Cushing’s disease, are associated with depression (Sonino et al., 1998). Within neuroinflammatory diseases such as multiple sclerosis use of high dose glucocorticoid treatment is thought to reduce BBB permeability and therefore reduce the flow of inflammatory mediators from the blood into the CNS (Miller et al., 1992). Anti-glucocorticoid treatments have also been used with some benefit in the treatment of MDD in patients with high serum glucocorticoids (Lombardo et al., 2019). Therefore, increases in circulating glucocorticoids in MDD, although not measured in this study, could potentially explain the reduction in BBB permeability demonstrated and therefore changes in BBB permeability could explain the mechanism of their actions.

It is also worth noting that TSPO ligands have molecular weight similar to that of other lipid-soluble molecules like antidepressants such as citalopram (∼300 g/mol) (Frisk-Holmberg and van der Kleijn, 1972) and thus it is interesting to consider the possibility that reduced small molecule perfusion into the brain could be a mechanism of drug resistance. In fact in those instances where MDD cohorts demonstrate increases in BBB porosity, drug response seems associated with increased levels of the marker S100β (Ambree et al., 2015; Arolt et al., 2003) although this is debated (Jang et al., 2008).

The observation of reduced transfer across the choroid plexus into CSF also supports our proposed model; amongst its various functions, the choroid plexus regulates the access for chemical substances from blood to the CSF playing a key role in neurotransmitter volumetric transmission (Skipor and Thiery, 2008). Molecular and gene-expression analysis of choroid plexus gene expressions in MDD has suggested alterations to calcium signalling as well as to immune activity in MDD (Devorak et al., 2015; Turner et al., 2014).

Finally, it is also of note the significant negative correlation between reduced CSF tracer exchange in ventricles and increased childhood trauma scores observed in the secondary analysis; this correlation did not survive multiple comparison correction but it is suggestive that early life events may translate into BBB porosity becoming a trait in the following years, most likely because of an increased inflammatory state (Baumeister et al., 2016), and become a risk factor for adult depression. Further investigations on the issue are needed in a much larger dataset.

### Methodological Issues

To measures the perfusion of the tracers from blood to CSF we focused on the amount of radioactivity percolating into the lateral ventricles. Generally, the source of this radioactivity is both the fraction of the radiotracer in parenchyma flowing in the interstitial spaces and then across the ependymal cells that line the ventricular spaces and the radiotracer flowing directly from blood through the choroid-plexus (Schubert et al., 2019). Kinetic modelling of the [11C]PBR28 data evidenced that in the case of TSPO tracers, likely due to their poor tissue extraction, radioactivity in the lateral ventricles is predominantly sourced from blood.

Given the variability across subjects in the BIODEP dataset as well as the un-availability of arterial blood data for [11C]PK11195 study, we then turned to another dataset where inflammation was experimentally controlled via IFN-α challenge in normal volunteers and measured with [11C]PBR28 with arterial sampling. Here we also observed a strong inverse linear relationship between peripheral CRP levels and radioactivity in ventricles measured both with SUVR and via kinetic modelling through the blood to lateral ventricle CSF transport. In these data as in the previous set, the effect was amplified non-linearly for abnormal CRP levels. Although we do not know whether CRP is directly inducing the observed reduction in tracer delivery, this modulation, that was consistently observed across the two datasets, possibly indicates a threshold effect with barriers becoming less permeable for inflammatory states corresponding to CRP levels greater than 3-5 mg/L.

In the [11C]PBR28 dataset, the same association between CRP and reduction in tracer delivery to the lateral ventricles was observed in the blood-to-brain transfer constant *K*_*1*_. This also suggests that in the BIODEP dataset the SUVR for the lateral ventricles may actually underestimate the reduction of delivery into those spaces given that is likely that the delivery to parenchyma was also reduced; this would explain the lack of group difference between controls and patients. Note that an alternative explanation is that the observed associations are confounded by changes in the free fraction of the radiotracer in plasma. Lockart et al., (Lockhart et al., 2003) demonstrated an affinity of PK11195, a quinoline, to the reactant alpha1-acid glycoprotein which, although not measured in these two datasets, is also likely to be positively associated with peripheral inflammatory states. There was indeed an association between tracer delivery into brain and CSF and plasma free fractions of [11C]PBR28, the latter being highly collinear with CRP levels. However, the range of changes in free fractions did not explain a significant portion of the variability of blood-to-tissue transport (as measured by *K_1_*) either in CSF or in GM.

The inverse association between TSPO tracer perfusion and peripheral inflammatory status indexed by CRP may explain why brain TSPO density quantified with a plasma input function has shown reductions or no differences in MDD cases versus HC as the parameter used, the volume of distribution, is dependent on the target TSPO density but also on the perfusion of the tracer into tissue. Reference tissue approaches to quantification, that are not dependent on plasma-to-tissue transfer, may be more suitable for the quantification task in these cohorts (Marques et al., 2019).

### Limitations

The use of PET tracer percolation into lateral ventricle is an indirect way to quantify CSF dynamics and suffers from the low PET signal measurable in this anatomical area. However, when we tested the reproducibility of our metrics (both SUVR and AUC30-60) in a test-retest [11C]PBR28 brain PET dataset of 5 patient with mild AD (Nair et al., 2016) we found good performances: for SUVR parameter the test-retest absolute variability was 8%±4%, corresponding to an intra-class correlation coefficient (ICC) = 0.97, while for AUC30-60 test-retest absolute variability was 12%±7% (ICC = 0.86). These numbers are in line with those obtained for SUV in whole GM for which absolute variability was 6%±3% and ICC = 0.94 (Nair et al., 2016).

## Conclusion

We have demonstrated a strong association between (increased) peripheral inflammation indexed by CRP and (reduced) blood-to-brain and blood-to-CSF transfer of TSPO-PET radiotracers. These results implicate a mechanism of reduced permeability of the BBB and BCSFB that may harmful if protracted by chronic inflammatory states, as well as explain the difficulties in the quantification of these radiotracers in the brain when used in subjects with inflammatory conditions. This mechanism should be further tested with radiomolecules tailored to assess specifically BBB porosity in these clinical cohorts and explore how altered BBB permeability could affect the passage of antidepressant medication SSRIs such as citalopram, which have similar molecular weights to these radiotracers.

## Data Availability

The datasets generated during and/or analysed during the current study are available from the corresponding author on reasonable request and upon approval from the research consortia. Dataset 1 is part of "Biomarkers in Depression (BIODEP) study, NIMA consortium, https://www.neuroimmunology.org.uk/. Dataset 2 is part of "BRC-FLAME study, Maudsley NIHR BRC (Neuroimaging theme), https://www.maudsleybrc.nihr.ac.uk

https://www.maudsleybrc.nihr.ac.uk

## Acknowledgements and Disclosures

The BIODEP study (dataset 1) was sponsored by the Cambridgeshire and Peterborough NHS Foundation Trust and the University of Cambridge, and funded by a strategic award from the Wellcome Trust (104025) in partnership with Janssen, GlaxoSmithKline, Lundbeck and Pfizer. Recruitment of participants was supported by the National Institute of Health Research (NIHR) Clinical Research Network: Kent, Surrey and Sussex & Eastern. Additional funding was provided by the National Institute for Health Research (NIHR) Biomedical Research Centre at South London and Maudsley NHS Foundation Trust and King’s College London, and by the NIHR Cambridge Biomedical Research Centre (Mental Health). The FLAME study (dataset 2) was sponsored by the NIHR Biomedical Research Centre at South London and Maudsley NHS Foundation Trust and King’s College London in partnership whit Janssen Pharmaceutical Companies of Johnson&Johnson. We would like to gratefully thank all study participants, research teams and laboratory staff, without whom this research would not have been possible.

## Compliance with ethical standards

All procedures performed in studies involving human participants were in accordance with the ethical standards of the institutional and/or national research committee and with the 1964 Helsinki declaration and its later amendments or comparable ethical standards. Written informed consent was obtained from all individual participants included in the studies. The BIODEP study was approved by the NRES Committee East of England Cambridge Central (REC reference:15/EE/0092) and the UK Administration of Radioactive Substances Advisory Committee. The FLAME study was approved by the Queen Square London Ethical committee (REC reference. 16/LO/1520)

